# Healthcare strain and intensive care during the COVID-19 outbreak in the Lombardy region: a retrospective observational study on 43,538 hospitalized patients

**DOI:** 10.1101/2020.11.06.20149690

**Authors:** Filippo Trentini, Valentina Marziano, Giorgio Guzzetta, Marcello Tirani, Danilo Cereda, Piero Poletti, Raffaella Piccarreta, Antonio Barone, Giuseppe Preziosi, Fabio Arduini, Petra Giulia Della Valle, Alberto Zanella, Francesca Grosso, Gabriele del Castillo, Ambra Castrofino, Giacomo Grasselli, Alessia Melegaro, Alessandra Piatti, Aida Andreassi, Maria Gramegna, Marco Ajelli, Stefano Merler

**Author notes:** Correspondence to: Dr. Filippo Trentini, Center for Information Technology, Bruno Kessler Foundation, Via Sommarive, 18, 38123 Trento, Italy and Dr. Marcello Tirani, Directorate General for Health of the Lombardy Region, Piazza Città di Lombardia 1, 20124 Milano, Italy.

## Abstract

**Background:** During the spring of 2020, the SARS-CoV-2 epidemic has caused significant resource strain in hospitals of Lombardy, Italy, with the demand for intensive care beds for COVID-19 patients exceeding the overall pre-crisis capacity. In this study, we evaluate the effect of healthcare strain on ICU admission and survival.

**Methods:** We used data on 43,538 patients admitted to a hospital in the region between February 20 and July 12, 2020, of which 3,993 (9.2%) were admitted to an ICU. We applied logistic regression to model the probability of being admitted to an ICU and the probability of survival among ICU patients. Negative binomial regressions were used to model the time between hospital and ICU admission and the length of stay in ICU.

**Results:** During the period of highest hospital strain (March 16 – April 22), individuals older than 70 years had a significantly lower probability of being admitted to an ICU and significantly longer times between hospital and ICU admission, indicating elective admission due to constrained resources. Healthcare strain did not have a clear effect on mortality, with the overall proportion of deaths declining from 52.1% (95%CI 49.8-54.5) for ICU patients admitted to the hospital before March 16, to 43.4% (95%CI 41.5-45.6) between March 16 and April 22, to 27.6% (95%CI 20.0-35.2) after April 22.

**Conclusions:** These data demonstrate and quantify the adoption of elective admission to ICUs during the peak phase of the SARS-CoV-2 epidemic in Lombardy. However, we show that for patients admitted to ICUs, clinical outcomes progressively improved despite the saturation of healthcare resources.

## INTRODUCTION

Italy was the first country in the Western hemisphere to be affected by a widespread epidemic of SARS-CoV-2 [1,2], and still has one of the highest cumulative burden of COVID-19 hospitalizations and deaths worldwide [3]. Lombardy, in particular, was the first and by far the hardest-hit Italian region, accounting alone for over half of COVID-19 hospital admissions in Italy [4], despite having about one sixth of the country’s population. The explosive spread of SARS-CoV-2 in the region, coupled with the high COVID-19 morbidity, threatened to collapse even one of the most advanced health systems in the country and has resulted in the rapid adoption of unprecedented control measures. Despite a rate of critical care beds per inhabitant above the European average [5] and drastic action, culminated in the national lockdown of March 11, hospitals of Lombardy were put under severe strain. By the end of March 2020, the bed occupancy due to COVID-19 in hospitals and Intensive Care units (ICUs) in the region was 11,883 and 1,324 respectively. For comparison, the pre-crisis total ICU capacity was 720 beds [6]. The rapid saturation of hospital capacity was predicted by mathematical models in the early phase of the epidemic [7], prompting the emergency expansion of COVID-19 dedicated ICU and hospital beds [8], similarly to what previously experienced during the epidemic in Wuhan [9]. In this study, we provide results on the impact of saturated healthcare resources on the probability and timing of admission to ICU, as well as on the survival of ICU patients, during the COVID-19 epidemic in Lombardy. We considered for this purpose data on all of the 43,538 patients with a COVID-19 diagnosis, admitted to hospitals in the region between February 21 and July 12, 2020.

## METHODS

Our study is based on retrospective data collected on the complete set of patients referred to the Regional Coordinating Center of Lombardy with a laboratory-confirmed SARS-CoV-2 infection and subsequently admitted to one of the 73 hospitals in the network between February 20 and July 12, 2020. As of July 12, 2020, the registry of patients hospitalized with a SARS-CoV-2 diagnosis in Lombardy includes 46,554 cases [4] over a population of 10,900,000 inhabitants. Laboratory confirmation of SARS-CoV-2 was defined as a positive result of real-time Reverse Transcriptase–Polymerase Chain Reaction (RT-PCR) assay of nasal and pharyngeal swabs and, in selected cases, confirmation with RT-PCR assay from lower respiratory tract aspirates.

The registry was specifically created for the SARS-CoV-2 epidemic through the surveillance information system of the Lombardy region. Data entry was performed either by the Local Health Autority (Agenzie di Tutela della Salute, ATS) or by health professionals (nurses or doctors) within hospitals. Once the data were verified by the ATS in charge, they were transferred to the regional information system. The institutional ethics board of Fondazione IRCCS Ca’ Granda Ospedale Maggiore Policlinico in Milan approved this study and due to the nature of retrospective chart review, waived the need for informed consent from individual patients.

### Study population

The analysis is focused on all patients who were admitted with COVID-19 after the onset of symptoms in Lombardy between February 21 and July 12, 2020, representing 93.9% of the total number of patients with a SARS-CoV-2 diagnosis (see the flowchart in Figure S1 in Appendix).

The remainder represented cases who had been admitted before the onset of symptoms for reasons different than COVID-19.

Data reported in this study were collected at hospital admission (age, sex, province of residence, date of symptom onset, date of SARS-CoV-2 diagnosis, date of admission) and throughout the course of the patient’s stay (date of admission and discharge from ICUs, if any, date of discharge from hospital, clinical outcome, i.e. recovery or death).

To address potential bias due to data entry errors, we excluded patients for whom we found inconsistencies in the different dates (163 patients, 0.37% of the dataset, see the flowchart in Figure S1 in Appendix), and the final dataset includes 43,538 patients.

Age was treated as a categorical variable and divided into 4 classes (0-55y, 55-69y, 70-79y and >79y) based on quartiles of the age distribution in the sample. Available data on comorbidities were aggregated by different types of clinical conditions: cardiovascular diseases include hypertension, hypercholesterolemia, myocardiopathy, heart failure, ischemic and valve cardiopathy, arterial and venous vasculopathy; respiratory diseases include respiratory insufficiency, COPD and asthma; metabolic disorders include all types of diabetes and thyroid disorders. We divided the period of hospitalization into three sub-periods: i) period 1, from February 21 to March 15, when the ICU occupancy was increasing but remained below 800 beds (exponential phase of the epidemic, see Figure S2); ii) period 2, from March 16 to April 22 when the ICU occupancy ranged between 800 and 1330 beds (period of maximum strain of the healthcare system in Lombardy); iii) period 3, from April 17 to July 12 2020, when the ICU occupancy fell again below the threshold of 800 beds (declining phase of the epidemic). Patients were followed up until the end of available data (July 12).

### Statistical analysis

The outcomes of our descriptive analyses are: the probability of being admitted to an ICU, the probability of death among ICU patients, the time from symptom onset to either hospital admission or clinical outcome, the time from hospital admission to either death or discharge from the hospital (length of stay in hospital), the time between hospitalization and admission in ICU, and the time from admission to an ICU to either death or discharge from the ICU (length of stay in ICU). Age and times between events are presented as median and interquartile range (IQR), while binary variables, e.g. admission in ICU and fatal outcome, are summarized by sample proportions and the corresponding 95% confidence intervals (CI).

When comparing two groups, we used: two-sided t-tests to assess the statistical significance of observed differences in length of ICU stay and in time from hospitalization to ICU admission; and tests on proportions to assess the statistical significance of the observed differences in the probability of ICU admission and in the probability of death. To assess differences across multiple groups, we used one-way ANOVA, followed by post-hoc Tukey test.

The outcomes of our multivariate analyses are the probability of being admitted in ICU, the probability of death among ICU patients, the time from hospitalization to ICU admission and the length of stay in ICU. Logistic regression models were used to estimate adjusted odds ratios (OR) and to investigate the effect of the period of hospitalization on the probability of being admitted to ICU, using age, sex, comorbidities and province of residence of patients as confounders.

Negative binomial regressions were used to estimate adjusted Incidence Rate Ratios (IRR) and to investigate the effect of the period of hospitalization on the time between hospitalization and ICU admission using age, sex, comorbidities, province of residence and the clinical outcome as confounders.

In all models, an interaction term between period and age was added to account for differences in estimates across different subgroups of patients. Negative binomial regression was preferred to Poisson regression based on the likelihood ratio test. Finally, the statistical significance of the parameters of the logistic and of the negative binomial regressions was assessed through the Wald test.

Statistical analysis was performed using R version 3.6.2, and packages “boot”, “MASS”, “multcomp”,”lmtest” and “aod”. Stratum-specific odds ratio were computed using STATA.

Statistical significance was defined as p-value < □0.05.

## RESULTS

### Descriptive analyses

Among the 43,538 patients hospitalized within July 12, 2020, 3,993 (9.2%) were admitted to an ICU. The median age of patients admitted to hospitals was 68 years (IQR 55-79), and the majority of patients were male (59.6%, Table 1). Among patients admitted to ICUs, the median age was 63 (IQR 56-70) and 78.8% were male.

**Table 1.**
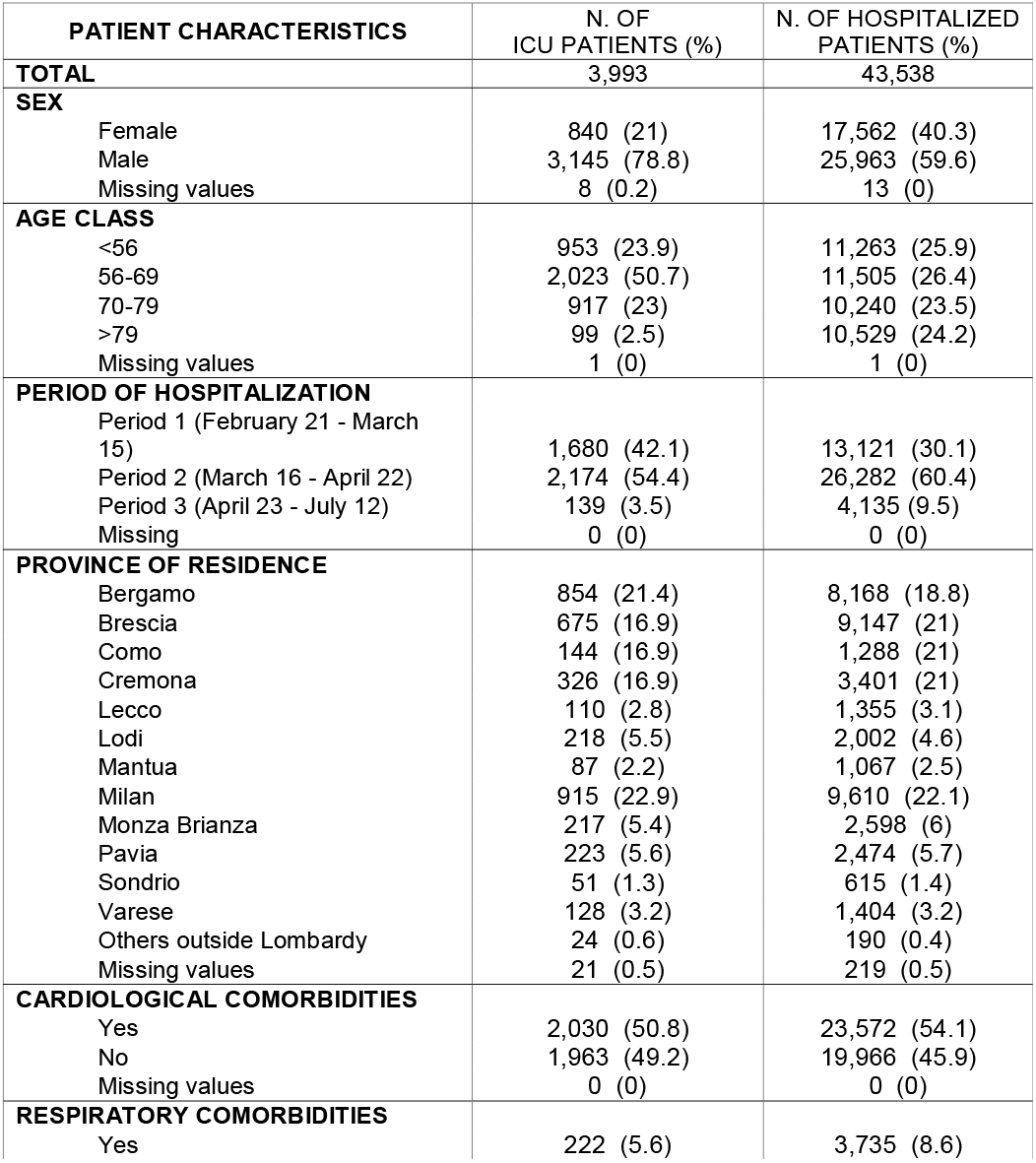
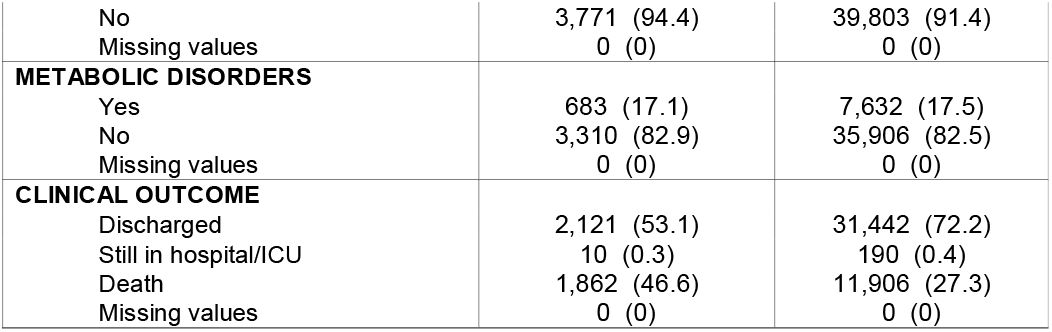
Characteristics of patients hospitalized and admitted to an ICU due to COVID-19 symptoms between February 21 and July 12, 2020, Lombardy region, Italy.

| **PATIENT CHARACTERISTICS** | **N. OF**<br>**ICU PATIENTS (%)** | **N. OF HOSPITALIZED**<br>**PATIENTS (%)** |
| --- | --- | --- |
| **TOTAL** | 3,993 | 43,538 |
| **SEX** |  |  |
| Female | 840 (21) | 17,562 (40.3) |
| Male | 3,145 (78.8) | 25,963 (59.6) |
| Missing values | 8 (0.2) | 13 (0) |
| **AGE CLASS** |  |  |
| <56 | 953 (23.9) | 11,263 (25.9) |
| 56-69 | 2,023 (50.7) | 11,505 (26.4) |
| 70-79 | 917 (23) | 10,240 (23.5) |
| >79 | 99 (2.5) | 10,529 (24.2) |
| Missing values | 1 (0) | 1 (0) |
| **PERIOD OF HOSPITALIZATION** |  |  |
| Period 1 (February 21 - March 15) | 1,680 (42.1) | 13,121 (30.1) |
| Period 2 (March 16 - April 22) | 2,174 (54.4) | 26,282 (60.4) |
| Period 3 (April 23 - July 12) | 139 (3.5) | 4,135 (9.5) |
| Missing | 0 (0) | 0 (0) |
| **PROVINCE OF RESIDENCE** |  |  |
| Bergamo | 854 (21.4) | 8,168 (18.8) |
| Brescia | 675 (16.9) | 9,147 (21) |
| Como | 144 (16.9) | 1,288 (21) |
| Cremona | 326 (16.9) | 3,401 (21) |
| Lecco | 110 (2.8) | 1,355 (3.1) |
| Lodi | 218 (5.5) | 2,002 (4.6) |
| Mantua | 87 (2.2) | 1,067 (2.5) |
| Milan | 915 (22.9) | 9,610 (22.1) |
| Monza Brianza | 217 (5.4) | 2,598 (6) |
| Pavia | 223 (5.6) | 2,474 (5.7) |
| Sondrio | 51 (1.3) | 615 (1.4) |
| Varese | 128 (3.2) | 1,404 (3.2) |
| Others outside Lombardy | 24 (0.6) | 190 (0.4) |
| Missing values | 21 (0.5) | 219 (0.5) |
| **CARDIOLOGICAL COMORBIDITIES** |  |  |
| Yes | 2,030 (50.8) | 23,572 (54.1) |
| No | 1,963 (49.2) | 19,966 (45.9) |
| Missing values | 0 (0) | 0 (0) |
| **RESPIRATORY COMORBIDITIES** |  |  |
| Yes | 222 (5.6) | 3,735 (8.6) |
| No | 3,771 (94.4) | 39,803 (91.4) |
| Missing values | 0 (0) | 0 (0) |
| **METABOLIC DISORDERS** |  |  |
| Yes | 683 (17.1) | 7,632 (17.5) |
| No | 3,310 (82.9) | 35,906 (82.5) |
| Missing values | 0 (0) | 0 (0) |
| **CLINICAL OUTCOME** |  |  |
| Discharged | 2,121 (53.1) | 31,442 (72.2) |
| Still in hospital/ICU | 10 (0.3) | 190 (0.4) |
| Death | 1,862 (46.6) | 11,906 (27.3) |
| Missing values | 0 (0) | 0 (0) |

There were 11,906 (27.3%) deaths among hospitalized patients, of which 1,862 among patients admitted to an ICU (46.6% of all ICU admissions). The majority of hospital and ICU admissions occurred during Period 2 (March 16 – April 22), corresponding to the highest healthcare strain in Lombardy.

The median time between symptoms and hospital admission among ICU patients was 6 days (IQR: 3-9, Table 2); the median time from hospitalization to ICU admission was 3 days (IQR: 0-6). Among ICU patients, those with a fatal outcome died after a median time of 15 (IQR: 9-24) days after hospital admission, while survivors were discharged from hospital after a median time of 38 (IQR: 24-55) days. The median length of stay in ICU for all patients irrespective of their clinical outcome was 11 days (IQR: 6-19). The length of stay in ICU was shorter for non-survivors than for survivors (median 10 vs 12 days; two-sided t-test on difference in means p-value<0.001).

**Table 2.**
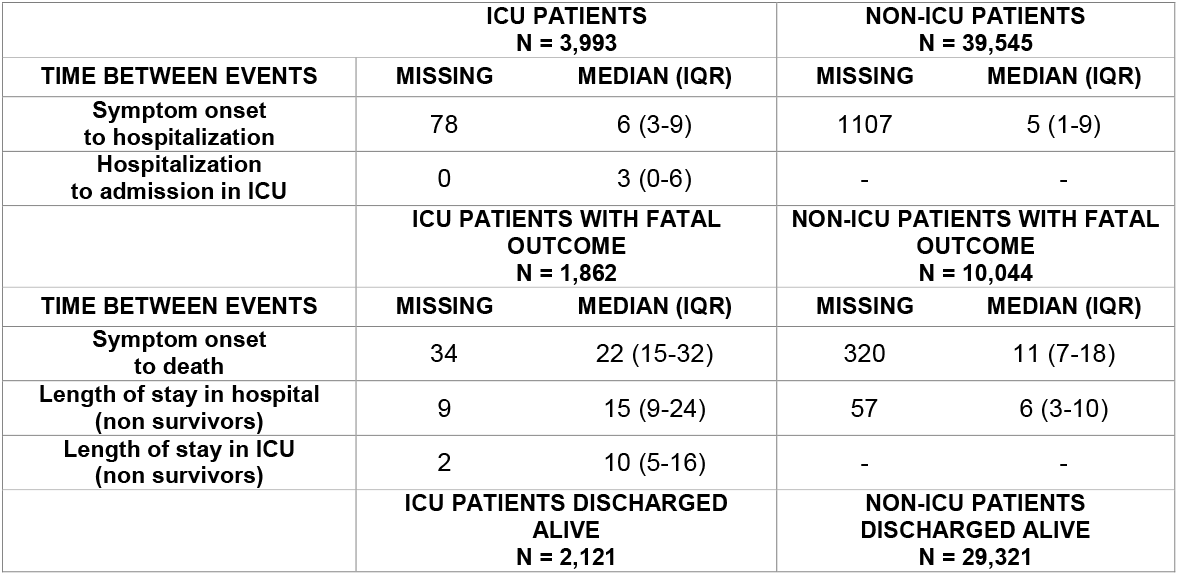
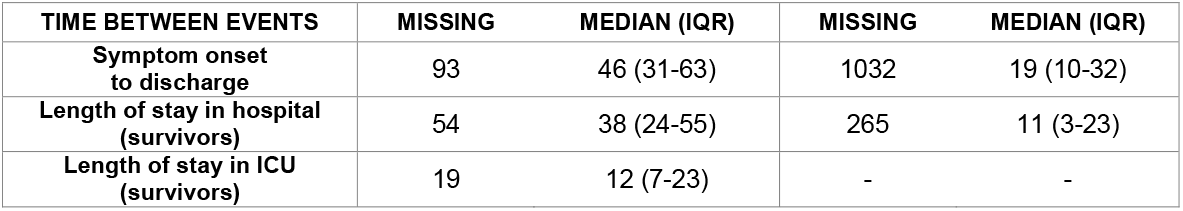
Key times-to-events for hospital admitted COVID-19 patients in Lombardy.

### Probability of ICU admission

The proportion of hospitalized patients that were admitted to an ICU shows an overall decreasing trend over the course of the epidemic (Figure 1-A), from 12.8% (95%CI: 12.2-13.4%) in the first period, to 8.2% (95%CI: 7.9-8.6%) in the second period, to 3.4% (95%CI: 2.8-4.0% in the third period; post-hoc Tukey test p-values on the differences in means <0.001). However, the ICU admission rates for patients aged above 70 declined more briskly during the month of March, when the availability of ICU beds was at its minimum, and then recovered partially in the following weeks.

**Figure 1.**
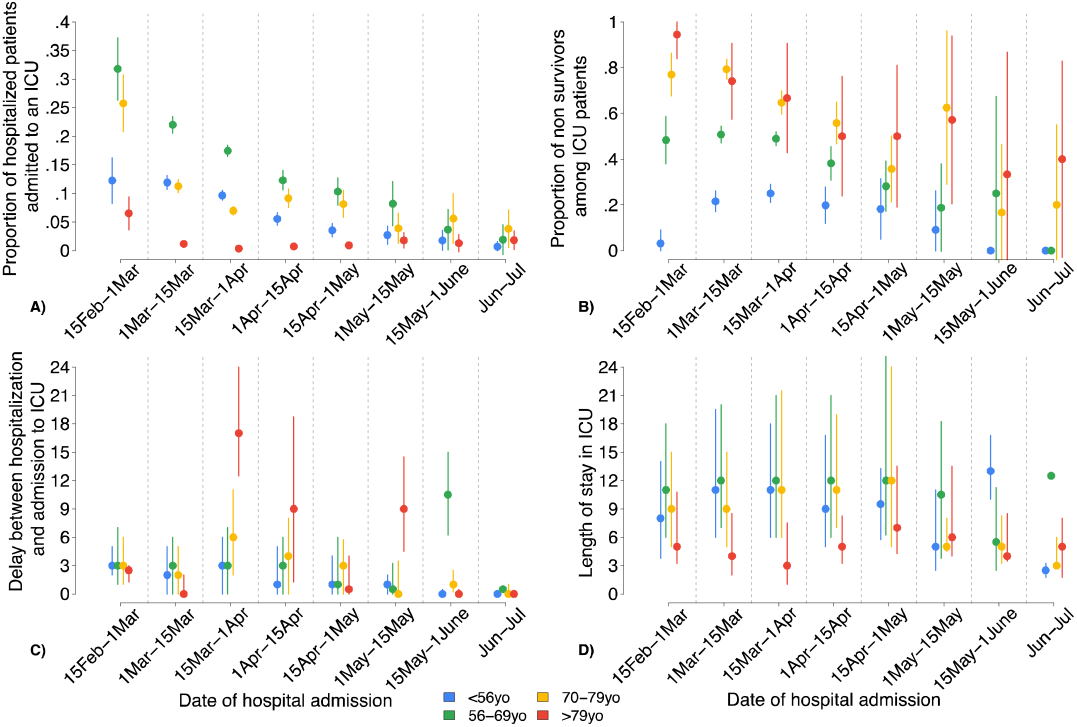
Average proportion of patients admitted to ICU among hospitalized patients (panel A) and average proportion of non-survivors among ICU patients (panel B), by date of hospital admission and age class; dots represent sample means and lines represents 95% CIs. Time between hospital and ICU admission (panel C) and length of stay in ICU (panel D), by date of hospital admission and age class; dots represent medians and lines represents IQRs.

This pattern is confirmed by the results of the regression analysis on the probability of being admitted in ICU, which showed a significant interaction between age and period of hospital admission (Table 3). This implies that the stratum-specific odds of being admitted in ICU among patients aged below 70 declined continuously from the first to the third period (in period 1 to 3 respectively 1.636, 1, 0.211; see Table 3), while those of patients aged 70-79 did not decrease in the third period and those of patients above 80 years of age were lowest in the second period (in period 1 to 3 respectively 2.61, 1, 4.76; see Table 3). The multivariate analysis on the probability of admission to an ICU also indicates that female patients, survivors and patients with comorbidities were overall less likely to be admitted in ICU (Table 3). In addition, we found that the odds of being admitted to an ICU increased by about 0.4% (95%CI −0.1-0.9%, Wald test p-value = 0.082) with every additional one-day increase in the delay between symptom onset and hospital admission.

**Table 3.**
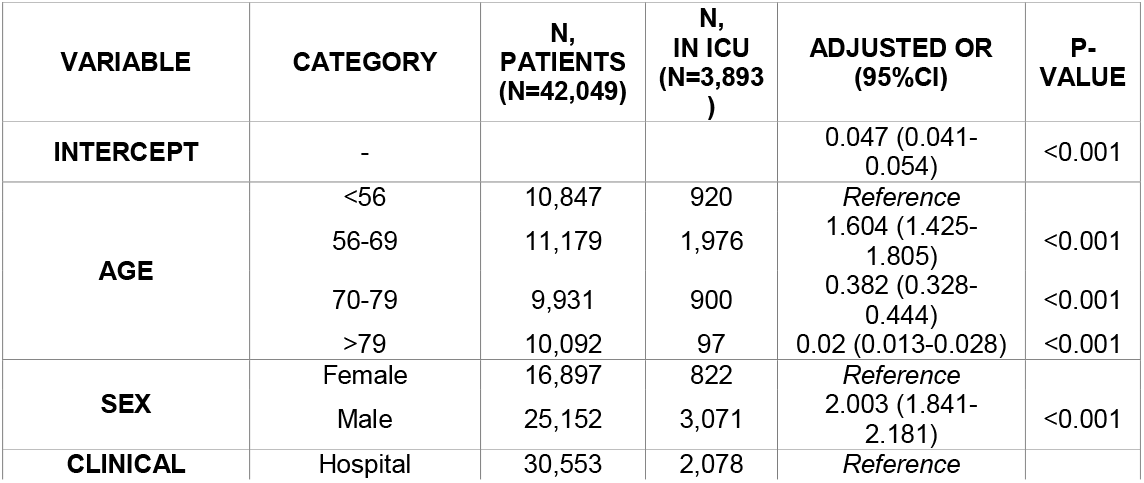
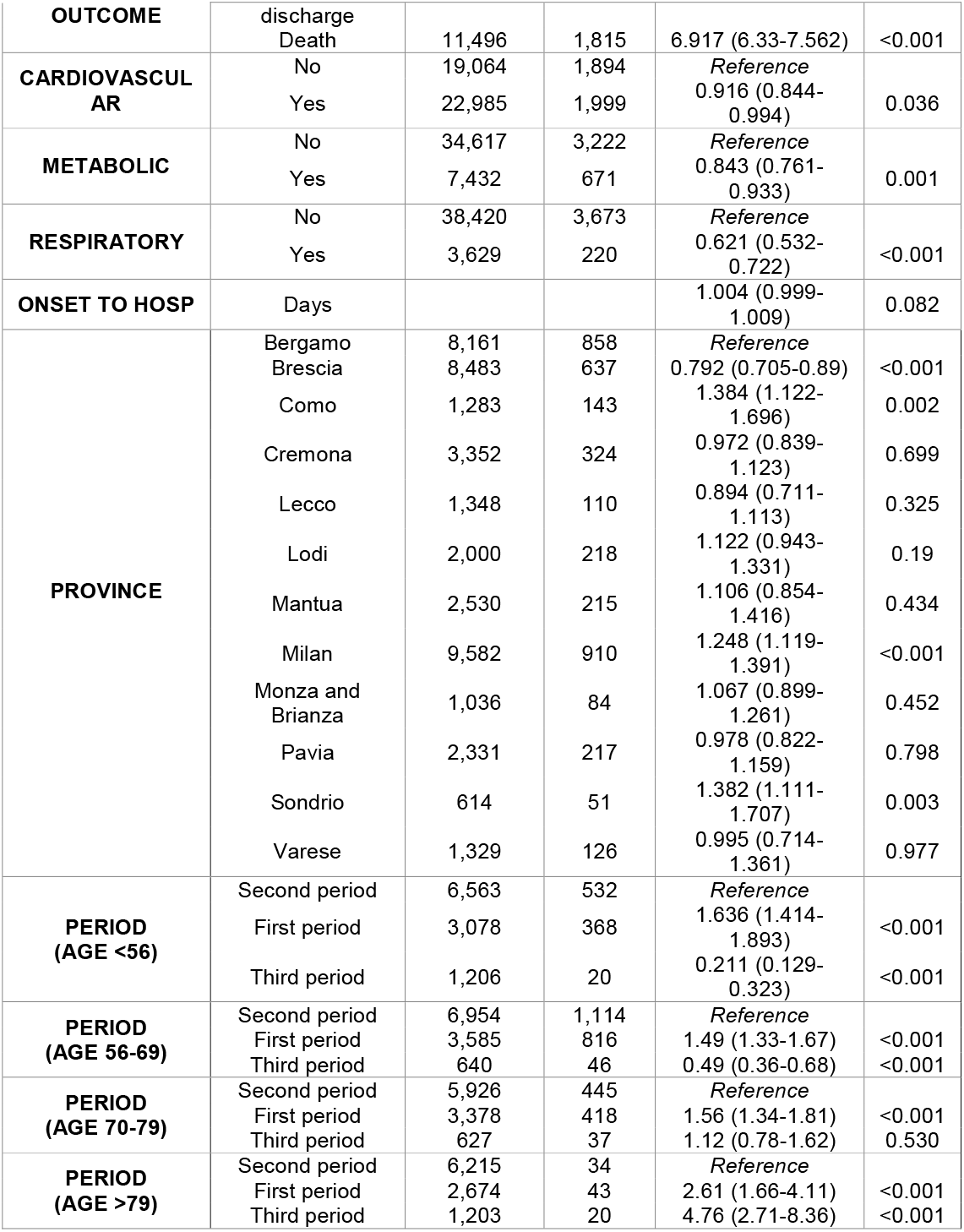
Adjusted OR and stratum-specific OR of being admitted in ICU and 95% confidence intervals. Interaction terms and relative 95%CI are reported in Table S1 of the Appendix.

| VARIABLE | CATEGORY | N,<br>PATIENTS<br>(N=42,049) | N,<br>IN ICU<br>(N=3,893) | ADJUSTED OR<br>(95%CI) | P-VALUE |
| --- | --- | --- | --- | --- | --- |
| INTERCEPT | - |  |  | 0.047 (0.041-0.054) | <0.001 |
| AGE | <56 | 10,847 | 920 | *Reference* |  |
|  | 56-69 | 11,179 | 1,976 | 1.604 (1.425-1.805) | <0.001 |
|  | 70-79 | 9,931 | 900 | 0.382 (0.328-0.444) | <0.001 |
|  | >79 | 10,092 | 97 | 0.02 (0.013-0.028) | <0.001 |
| SEX | Female | 16,897 | 822 | *Reference* |  |
|  | Male | 25,152 | 3,071 | 2.003 (1.841-2.181) | <0.001 |
| CLINICAL | Hospital | 30,553 | 2,078 | *Reference* |  |
| **OUTCOME** | discharge |  |  |  |  |
|  | Death | 11,496 | 1,815 | 6.917 (6.33-7.562) | <0.001 |
| **CARDIOVASCULAR** | No | 19,064 | 1,894 | *Reference* |  |
|  | Yes | 22,985 | 1,999 | 0.916 (0.844-0.994) | 0.036 |
| **METABOLIC** | No | 34,617 | 3,222 | *Reference* |  |
|  | Yes | 7,432 | 671 | 0.843 (0.761-0.933) | 0.001 |
| **RESPIRATORY** | No | 38,420 | 3,673 | *Reference* |  |
|  | Yes | 3,629 | 220 | 0.621 (0.532-0.722) | <0.001 |
| **ONSET TO HOSP** | Days |  |  | 1.004 (0.999-1.009) | 0.082 |
| **PROVINCE** | Bergamo | 8,161 | 858 | *Reference* |  |
|  | Brescia | 8,483 | 637 | 0.792 (0.705-0.89) | <0.001 |
|  | Como | 1,283 | 143 | 1.384 (1.122-1.696) | 0.002 |
|  | Cremona | 3,352 | 324 | 0.972 (0.839-1.123) | 0.699 |
|  | Lecco | 1,348 | 110 | 0.894 (0.711-1.113) | 0.325 |
|  | Lodi | 2,000 | 218 | 1.122 (0.943-1.331) | 0.19 |
|  | Mantua | 2,530 | 215 | 1.106 (0.854-1.416) | 0.434 |
|  | Milan | 9,582 | 910 | 1.248 (1.119-1.391) | <0.001 |
|  | Monza and Brianza | 1,036 | 84 | 1.067 (0.899-1.261) | 0.452 |
|  | Pavia | 2,331 | 217 | 0.978 (0.822-1.159) | 0.798 |
|  | Sondrio | 614 | 51 | 1.382 (1.111-1.707) | 0.003 |
|  | Varese | 1,329 | 126 | 0.995 (0.714-1.361) | 0.977 |
| **PERIOD (AGE <56)** | Second period | 6,563 | 532 | *Reference* |  |
|  | First period | 3,078 | 368 | 1.636 (1.414-1.893) | <0.001 |
|  | Third period | 1,206 | 20 | 0.211 (0.129-0.323) | <0.001 |
| **PERIOD (AGE 56-69)** | Second period | 6,954 | 1,114 | *Reference* |  |
|  | First period | 3,585 | 816 | 1.49 (1.33-1.67) | <0.001 |
|  | Third period | 640 | 46 | 0.49 (0.36-0.68) | <0.001 |
| **PERIOD (AGE 70-79)** | Second period | 5,926 | 445 | *Reference* |  |
|  | First period | 3,378 | 418 | 1.56 (1.34-1.81) | <0.001 |
|  | Third period | 627 | 37 | 1.12 (0.78-1.62) | 0.530 |
| **PERIOD (AGE >79)** | Second period | 6,215 | 34 | *Reference* |  |
|  | First period | 2,674 | 43 | 2.61 (1.66-4.11) | <0.001 |
|  | Third period | 1,203 | 20 | 4.76 (2.71-8.36) | <0.001 |

### Probability of death among ICU patients

Figure 1-B shows an increasing trend in the mortality of ICU patients below 60 years during the month of March, in striking contrast with the declining trend of mortality for patients older than 70 in the same month. The overall proportion of non-survivors among ICU patients decreased from 52.1% (95%CI 49.8-54.5) for patients hospitalized during period 1 to 43.4% (95%CI 41.5-45.6) and 27.6% (95%CI 20.0-35.2) for those admitted respectively during the second and third period (post-hoc Tukey test p-values on the differences in means <0.001). In a multivariate analysis, we found the interaction between period and age to be significant, with ICU patients above 70 years of age having a continuously decreasing risk of death from period 1 to period 3, and younger individuals having similar odds of dying over the three periods (see Table S3). The odds of dying decrease as the delay between hospitalization and ICU admission increases (OR 0.983, 95%CI: 0.974 −0.991, Table S3).

### Time between hospital and ICU admission

From patients aged 70 years and older, there was a brisk increase in the time between hospital and ICU admission starting from mid-March (Figure 1C). Considering all patients, the average time between hospital and ICU admission during period 2 was 1.25 days (95%CI 0.63-1.87) longer compared to period 1 and 2.14 days (95%CI 0.44-3.84) longer compared to period 3 (post-hoc Tukey test p-values <0.001 and 0.009 respectively); the difference between period 3 and period 1 was not significant (mean difference = 0.90, post-hoc Tukey test p-value=0.43). The interaction between age and period turned out to be significant in a negative binomial regression on the delay between hospitalization and admission to ICU (Table 4). The adjusted stratum-specific IRRs indicate that the time between hospital and ICU admission for patients aged 70 or older was significantly higher during the second period compared to the first and third, while it did not change significantly for younger patients (Table 4).

**Table 4.** Adjusted IRR and stratum-specific IRR of delay between hospitalization and admission in ICU and 95% confidence intervals. Interaction terms and relative 95%CI are reported in Table S2 of the Appendix.

| VARIABLE | CATEGORY | N. PATIENTS<br>(n=3,945) | Adjusted IRR<br>(95%CI) | P-VALUE |
| --- | --- | --- | --- | --- |
| INTERCEPT | - | - | 4.289 (3.647-5.059) | <0.001 |
| AGE | Age class <56 | 935 | Reference |  |
|  | Age class 56-69 | 1,999 | 1.207 (1.045-1.392) | 0.01 |
|  | Age class 70-79 | 913 | 1.803 (1.512-2.151) | <0.001 |
|  | Age class >79 | 98 | 3.321 (2.148-5.415) | <0.001 |
| SEX | Female | 833 | Reference |  |
|  | Male | 3,112 | 0.973 (0.875-1.08) | 0.61 |
| CLINICAL<br>OUTCOME | Hospital<br>discharge | 2,105 | Reference |  |
|  | Death | 1,840 | 0.827 (0.755-0.906) | <0.001 |
| CARDIOVASCU<br>LAR | No | 1,915 | Reference |  |
|  | Yes | 2,030 | 1.066 (0.97-1.17) | 0.18 |
| METABOLIC | No | 3,262 | Reference |  |
|  | Yes | 683 | 1.002 (0.891-1.129) | 0.976 |
| RESPIRATORY | No | 3,723 | Reference |  |
|  | Yes | 222 | 1.082 (0.903-1.307) | 0.402 |
| PROVINCE | Bergamo | 854 | Reference |  |
|  | Brescia | 675 | 1.362 (1.187-1.563) | <0.001 |
|  | Como | 144 | 1.037 (0.818-1.327) | 0.771 |
|  | Cremona | 325 | 1.234 (1.037-1.472) | 0.018 |
|  | Lecco | 110 | 1.245 (0.958-1.64) | 0.11 |
|  | Lodi | 218 | 0.875 (0.713-1.08) | 0.206 |
|  | Mantua | 217 | 0.951 (0.707-1.301) | 0.744 |
|  | Milan | 913 | 1.037 (0.911-1.18) | 0.583 |
|  | Monza and<br>Brianza | 87 | 1.158 (0.948-1.423) | 0.156 |
|  | Pavia | 223 | 1.412 (1.159-1.73) | 0.001 |
|  | Sondrio | 51 | 1.31 (1.025-1.695) | 0.035 |
|  | Varese | 128 | 0.676 (0.463-1.018) | 0.051 |
| PERIOD<br>(AGE <56) | Second period | 543 | Reference |  |
|  | First period | 372 | 0.86 (0.717-1.034) | 0.106 |
|  | Third period | 20 | 0.507 (0.274-1.012) | 0.059 |
| PERIOD<br>(AGE 56-69) | Second period | 1,128 | Reference |  |
|  | First period | 824 | 0.903 (0.798-1.022) | 0.108 |
|  | Third period | 47 | 0.803 (0.537-1.202) | 0.287 |
| PERIOD<br>(AGE 70-79) | Second period | 448 | Reference |  |
|  | First period | 426 | 0.607 (0.506-0.729) | <0.001 |
|  | Third period | 39 | 0.45 (0.269-0.670) | <0.001 |
| PERIOD<br>(AGE >79) | Second period | 34 | Reference |  |
|  | First period | 44 | 0.326 (0.179-0.593) | <0.001 |
|  | Third period | 20 | 0.339 (0.162-0.708) | 0.004 |

### Length of stay in ICU

The length of stay in ICU was shorter among patients aged 80 or more hospitalized between mid-March and mid-April compared to other periods but tended to be longer for younger patients (Figure 1-D). Considering all patients independently of their age, the average length of stay in ICU during period 2 was 1.28 days longer compared to period 1 and 3.91 days longer compared to period 3 (post-hoc Tukey test p-values 0.007 and 0.003 respectively). The difference between period 1 and period 3 was 2.63 days (post-hoc Tukey test p-value 0.07). The length of stay in ICU was significantly longer for survivors (Table S4).

## DISCUSSION

In the spring of 2020, the healthcare system of the Lombardy region was under intense pressure due to the SARS-CoV-2 epidemic, with hospital and ICU bed capacity saturated by large numbers of COVID-19 patients. In addition to the limited availability of beds, the strain on the healthcare system was aggravated by the shortage of hospital workforce, due to the large number of infections occurring among doctors, nurses and other healthcare professionals [7], and to precautionary quarantines needed to limit hospital transmission. In this work, we identify a clear age-specific effect of the healthcare strain on the dynamics of admission to ICU. In particular, individuals older than 70 years were less likely to be admitted to an ICU during the peak period of ICU occupation, and those who were admitted had a longer delay since hospital admission compared to periods of higher bed availability. The saturation of healthcare resources did not have a clear negative effect on the survival of patients admitted to ICUs. In particular, the overall proportion of non-survivors declined from about 52% in the first phase of exponential growth of the epidemic (until March 15, 2020), to 43% in the central period characterized by highest occupation of ICU and hospital beds (March 16-April 22), to 28% in the declining phase of the epidemic (April 23-July 12). In line with a previous study [10], we found that the chance of surviving increased for patients admitted later to ICU, probably reflecting a lower severity among such patients [10]. ICU patients older than 70 years also had lower odds of dying in the period of peak ICU occupancy with respect to the early epidemic period, and comparable to patients of the same age admitted during the declining phase (end of April – mid July). This trend is not matched in younger age classes and is suggestive of elective admission in ICU based on chances of survival for older patients. Elective admission is in accordance with recommendations provided to face scarcity of resources during the COVID-19 pandemic [11, 12, 13]. In general, interpretation of mortality trends for ICU patients over time is made complex by the superposition of multiple dynamic factors, such as the varying level of healthcare strain and the progressive improvement of clinical and pharmaceutical practices.

This work has some limitations. First, we defined the period of healthcare strain solely on the basis of ICU bed occupancy, largely overlapped with the temporal evolution of the hospital bed occupancy (Figure S2). Further data to better characterize different aspects of hospital strain, such as the availability of ventilators, healthcare staff, drugs used for COVID-19 therapy and personal protective equipment, were not available. In addition, we did not have information about inter-hospital referrals which might provide further insight in the dynamics of healthcare strain for individual hospitals. The somewhat arbitrary threshold on the number of occupied ICU beds adopted for the definition of the three periods in our analysis may be a possible source of bias. This threshold was chosen on the basis of the pre-emergency ICU bed availability and in the attempt to obtain a good trade-off between balancing the number of hospitalized patients in the three periods and balancing the temporal duration of the periods. Second, we did not have more granular information on the individual therapeutic course and on the use of life support measures.

According to a recent report, about 87% of ICU patients in Lombardy received invasive mechanical ventilation, while the remaining were assisted with non-invasive respiratory support [14]. Third, information on individual comorbidities was coarsely represented by three macro categories (cardiovascular, respiratory and metabolic) that contained diverse conditions with a heterogeneous level of prognostic relevance. For this reason, the binary information on the presence or absence of a given type of comorbidity was only used as a confounder in the multivariate analyses.

Many studies have focused on risk factors associated with mortality [15,16,17,18] and ICU admission [18] among hospitalized patients. To the best of our knowledge, the effect of healthcare strain during the SARS-CoV-2 epidemic on ICU admission dynamics and survival of critical patients has not yet been investigated. Early observations have suggested an association between incidence levels of COVID-19 and the case-fatality ratios by comparing surveillance data in the general population from Wuhan, the province of Hubei and other provinces in China [19]. A more recent analysis has identified a positive association between the proportion of occupied ICU beds and the increase of COVID-19 deaths in the general population over the following 7 days, in a setting which was quite far from saturation (average ICU occupation 20%) [20].

## CONCLUSIONS

In this work, the use of complete hospitalization data from 43,558 COVID-19 patients in Lombardy, the largest and hardest-hit Italian region, allowed a detailed characterization of age-specific trends in intensive care management during the SARS-CoV-2 epidemic. In particular, we demonstrate and quantify the adoption of elective admission to ICUs during the peak phase of the epidemic. However, the mortality of patients admitted to an ICU progressively improved despite the constrained hospital resources. The data presented here on the probability and timing of admission to ICU, on the length of stay and on the survival of critical patients over the course of epidemic can be useful to inform disease management in other settings with saturated healthcare systems.

## Data Availability

Individual data used for the analyses will be made available in case of acceptance and published along with the manuscript.

## LIST OF ABBREVIATIONS

WHO: World Health Organization
IQR: Interquartile range
IRR: Incidence rate ratio
ICU: Intensive Care Unit
OR: Odds ratio

## Contributors

FT, VM, GGu, SM conceived and designed the study. FT, VM and RP performed the analysis. FT, VM, RP, GGu, PP, MA, SM contributed to data interpretation. MT, DC, AB, GP, FA, PDV, AZ, AP, GGr, AA, MG collected data. MT, DC verified all data. AM, RP and FT processed the data. FT, GGu wrote the first draft. All authors contributed critical revision of the manuscript and approved the final version of the manuscript.

## Acknowledgments

Not applicable.

