## Appendix for "Healthcare strain and intensive care during the COVID-19 outbreak in the Lombardy region: a retrospective observational study on 43,538 hospitalized patients"

Additional file 1

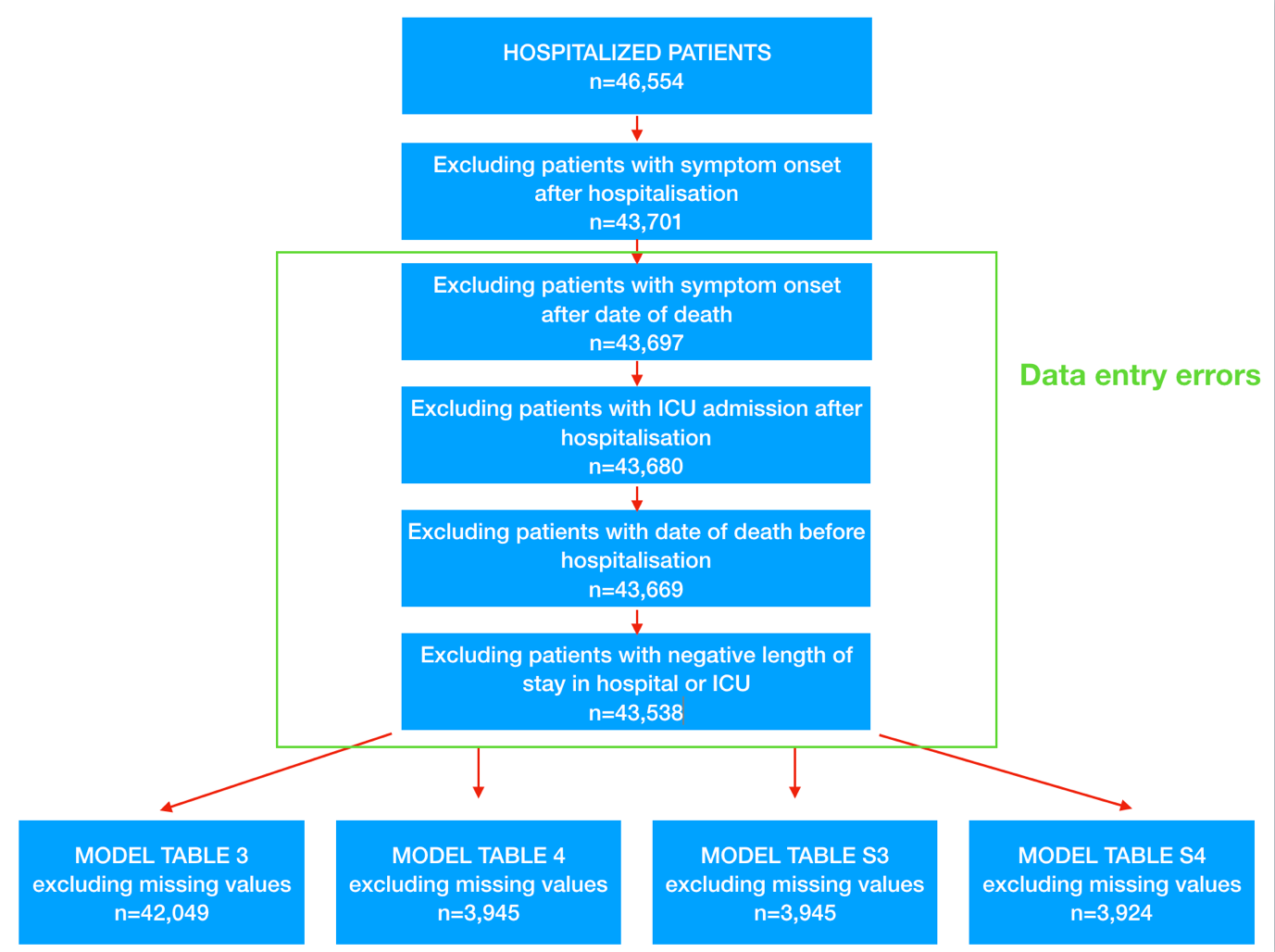

**Figure S1.** Flowchart with the sample sizes used for each analysis

*Hospital and ICU bed occupancy during the COVID-19 outbreak in Lombardy*

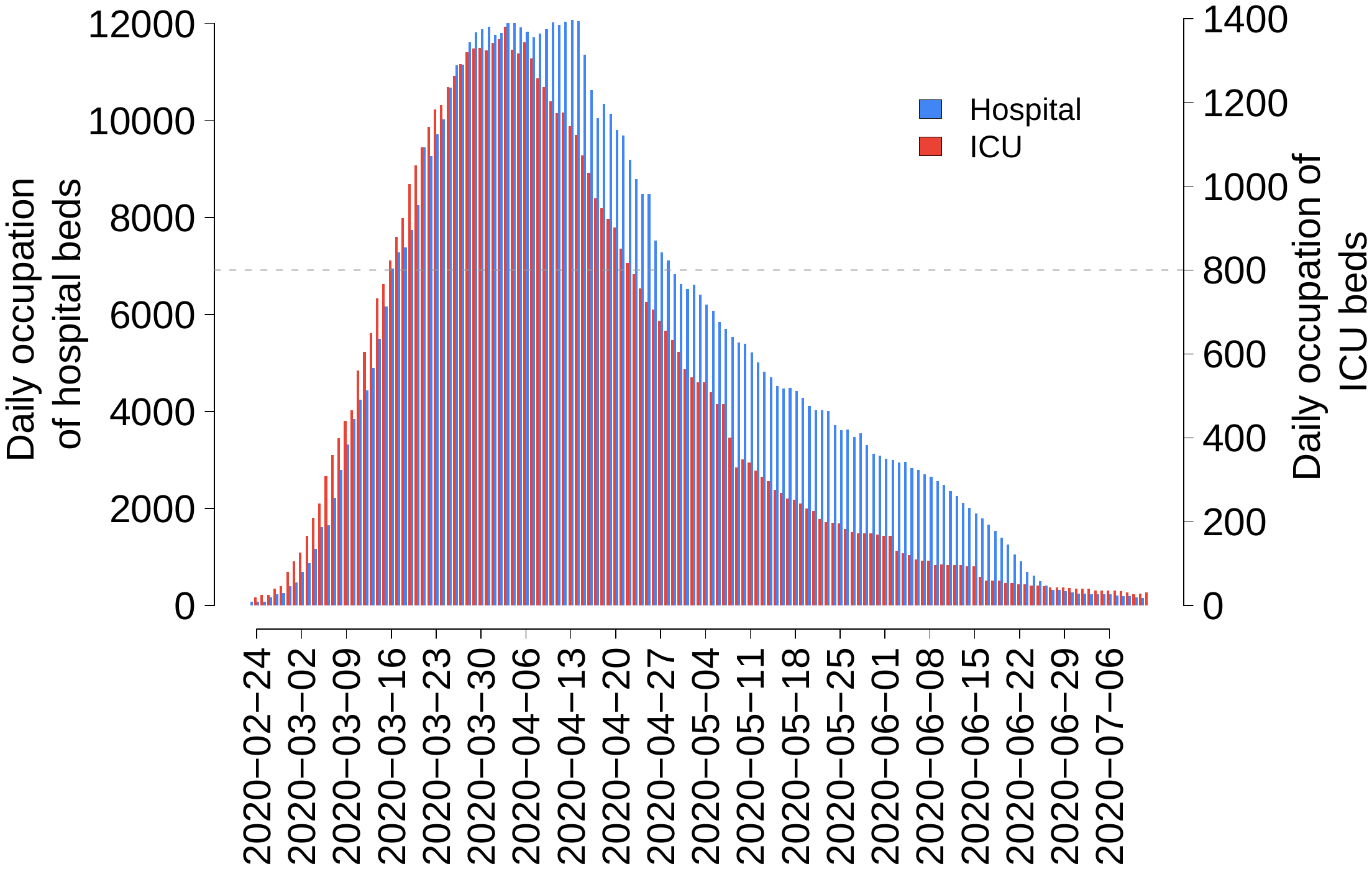

**Figure S2.** Number of daily occupied beds in hospital (blue) and ICU (red), between February 21 and July 12, 2020. The horizontal dashed grey line indicates the threshold on ICU beds adopted to define the three periods in the main analysis.

*Logistic regression on the probability of ICU admission*

Table S1 reports the adjusted ORs for the interaction term in the logistic regression model on the probability of ICU admission and integrates Table 3 in the main text. The stratum-specific odds ratios for being admitted in ICU in period P among patients in age class A, shown in Table 3, is given by the product of the OR of being admitted in ICU in the period P with respect to reference period P_r_ among patients in the reference age class A_r_, and the OR relative to the interaction term of age class A and period P. We consider P_r_ to be the second period and A_r_ to be individuals of age <56. For example, for P = period 3 and A = “> 79”, the two OR to be multiplied are respectively 0.211 (Table 3) and 22.6 (Table S1): hence the stratum-specific OR is 4.76 (Table 3).

**Table S1.** Estimated Interaction terms with 95% confidence intervals

| VARIABLE | CATEGORY | Adjusted OR (95%CI) | P-VALUE |
| --- | --- | --- | --- |
| Interaction  PERIOD * AGE | First period * age 56-69 | 0.91 (0.76-1.092) | 0.31 |
|  | First period * age 70-79 | 0.951 (0.774-1.169) | 0.632 |
|  | First period * age >79 | 1.593 (0.992-2.578) | 0.055 |
|  | Third period * age 56-69 | 2.326 (1.358-4.12) | 0.003 |
|  | Third period * age 70-79 | 5.332 (3.019-9.668) | <0.001 |
|  | Third period * age >79 | 22.6 (11-46.806) | <0.001 |

*Negative binomial regression on time between hospitalization and ICU admission*

Table S2 reports the adjusted IRRs for the interaction term in the negative binomial regression model on the time between hospitalization and ICU admission, integrating Table 4 in the main text.

**Table S2.** Estimated Interaction terms with 95% confidence intervals

| VARIABLE | CATEGORY | Adjusted IIR (95%CI) | P-VALUE |
| --- | --- | --- | --- |
| Interaction  PERIOD * AGE | First period * age 56-69 | 1.05 (0.843-1.306) | 0.661 |
|  | First period * age 70-79 | 0.706 (0.546-0.911) | 0.008 |
|  | First period * age >79 | 0.379 (0.201-0.706) | 0.002 |
|  | Third period * age 56-69 | 1.583 (0.72-3.335) | 0.231 |
|  | Third period * age 70-79 | 0.837 (0.371-1.821) | 0.657 |
|  | Third period * age >79 | 0.668 (0.248-1.784) | 0.417 |

*Logistic regression on probability of death among ICU patients*

A multivariate logistic regression on the probability of death among ICU patients suggests that older patients, male patients, patients with comorbidities and patients hospitalized earlier with respect to symptom onset are at higher risk of death (Table S3). Stratum-specific odds ratio of dying for patients over 70 decrease consistently during the three periods (Table S4). On the other hand, stratum-specific odds ratio of dying for patients under 70 years are not significantly different across the course of the epidemic.

**Table S3.** Model Estimates and 95% confidence intervals of OR and stratum-specific OR

| VARIABLE | CATEGORY | N. PATIENTS  (n=3,945) | n.  NON-SURVIVORS  (n=1,840) | ADJUSTED OR (95%CI) | P-VALUE |
| --- | --- | --- | --- | --- | --- |
| INTERCEPT | - | - |  | 0.209 (0.156-0.277) | <0.001 |
| AGE | Age class <56 | 935 | 204 |  |  |
|  | Age class 56-69 | 1,999 | 950 | 2.617 (2.068-3.326) | <0.001 |
|  | Age class 70-79 | 913 | 621 | 4.529 (3.401-6.06) | <0.001 |
|  | Age class >79 | 98 | 65 | 4.637 (2.21-10.003) | <0.001 |
| SEX | Female | 833 | 339 |  |  |
|  | Male | 3,112 | 1501 | 1.451 (1.225-1.72) | <0.001 |
| CARDIOLOGICAL COMORBIDITIES | No | 1,915 | 700 |  |  |
|  | Yes | 2,030 | 1140 | 1.494 (1.291-1.728) | <0.001 |
| METABOLIC COMORBIDITIES | No | 3,262 | 1443 |  |  |
|  | Yes | 683 | 397 | 1.285 (1.068-1.548) | 0.008 |
| RESPIRATORY COMORBIDITIES | No | 3,723 | 1721 |  |  |
|  | Yes | 222 | 119 | 1.143 (0.845-1.547) | 0.386 |
| TIME BETWEEN HOSPITAL AND ICU ADMISSION | Days |  |  | 0.983 (0.974-0.991) | <0.001 |
| Province | Bergamo | 854 | 388 |  |  |
|  | Brescia | 675 | 332 | 1.064 (0.855-1.324) | 0.58 |
|  | Como | 144 | 59 | 0.915 (0.62-1.345) | 0.653 |
|  | Cremona | 325 | 170 | 1.065 (0.805-1.409) | 0.659 |
|  | Lecco | 110 | 50 | 0.962 (0.625-1.477) | 0.861 |
|  | Lodi | 218 | 117 | 1.139 (0.818-1.586) | 0.442 |
|  | Mantua | 217 | 80 | 0.993 (0.608-1.612) | 0.976 |
|  | Milan | 913 | 394 | 0.967 (0.788-1.187) | 0.748 |
|  | Monza and Brianza | 87 | 39 | 0.603 (0.431-0.838) | 0.003 |
|  | Pavia | 223 | 133 | 1.967 (1.424-2.727) | <0.001 |
|  | Sondrio | 51 | 28 | 0.743 (0.49-1.117) | 0.156 |
|  | Varese | 128 | 50 | 1.11 (0.611-2.034) | 0.734 |
| Interaction PERIOD * AGE | First period * 56-69 | - |  | 1.414 (0.975-2.059) | 0.069 |
|  | First period * 70-79 | - |  | 2.916 (1.869-4.573) | <0.001 |
|  | First period * >79 | - |  | 3.274 (1.102-10.222) | 0.036 |
|  | Third period * 56-69 | - |  | 0.822 (0.184-5.832) | 0.816 |
|  | Third period * 70-79 | - |  | 1.198 (0.276-8.438) | 0.828 |
|  | Third period * >79 | - |  | 1.593 (0.274-13.362) | 0.627 |
| period  (AGE <56) | Second period | 543 | 127 | *Reference* |  |
|  | First period | 372 | 75 | 0.78 (0.56-1.08) | 0.139 |
|  | Third period | 20 | 2 | 0.37 (0.08-1.61) | 0.184 |
| period  (AGE 56-69) | Second period | 1,128 | 522 | *Reference* |  |
|  | First period | 824 | 418 | 1.10 (0.92-1.33) | 0.299 |
|  | Third period | 47 | 10 | 0.30 (0.15-0.62) | 0.001 |
| period  (AGE 70-79) | Second period | 448 | 269 | *Reference* |  |
|  | First period | 426 | 337 | 2.28 (1.67-3.10) | <0.001 |
|  | Third period | 39 | 15 | 0.44 (0.22-0.87) | 0.019 |
| period  (AGE >79) | Second period | 34 | 20 | *Reference* |  |
|  | First period | 44 | 36 | 2.56 (0.89-7.36) | 0.082 |
|  | Third period | 20 | 9 | 0.58 (0.18-1.85) | 0.359 |

*Negative binomial regression on length of ICU stay*

Table S4 reports the adjusted IRRs for the negative binomial regression model on the length of stay in ICU.

**Table S4.** Model Estimates and 95% confidence intervals of IRR *

| VARIABLE | CATEGORY | N. PATIENTS  (n=3,924) | ADJUSTED IRR (95%CI) | P-VALUE |
| --- | --- | --- | --- | --- |
| INTERCEPT | - |  | 15.19 (13.736-16.817) | <0.001 |
| AGE | Age class <56 | 929 |  |  |
|  | Age class 56-69 | 1,990 | 1.203 (1.103-1.31) | <0.001 |
|  | Age class 70-79 | 908 | 1.301 (1.169-1.449) | <0.001 |
|  | Age class >79 | 97 | 0.735 (0.549-1) | 0.043 |
| SEX | Female | 830 |  |  |
|  | Male | 3,094 | 1.019 (0.956-1.086) | 0.554 |
| CLINICAL OUTCOME | Survivor | 2,085 |  |  |
|  | Non-survivor | 1,839 | 0.689 (0.653-0.728) | <0.001 |
| CARDIO | No | 1,903 |  |  |
|  | Yes | 2,021 | 1.015 (0.96-1.073) | 0.614 |
| METABOLIC | No | 3,248 |  |  |
|  | Yes | 676 | 0.94 (0.877-1.01) | 0.092 |
| RESPIRATORY | No | 3,704 |  |  |
|  | Yes | 220 | 0.986 (0.881-1.106) | 0.803 |
| Province | Bergamo | 851 |  |  |
|  | Brescia | 673 | 0.956 (0.88-1.04) | 0.294 |
|  | Como | 143 | 0.812 (0.701-0.943) | 0.006 |
|  | Cremona | 325 | 0.843 (0.758-0.939) | 0.002 |
|  | Lecco | 109 | 1.226 (1.045-1.448) | 0.014 |
|  | Lodi | 218 | 0.959 (0.849-1.087) | 0.509 |
|  | Mantua | 216 | 0.909 (0.759-1.097) | 0.308 |
|  | Milan | 903 | 0.964 (0.892-1.042) | 0.355 |
|  | Monza and Brianza | 87 | 1.037 (0.918-1.174) | 0.562 |
|  | Pavia | 220 | 1.142 (1.012-1.291) | 0.033 |
|  | Sondrio | 51 | 1.221 (1.051-1.425) | 0.01 |
|  | Varese | 128 | 0.909 (0.723-1.157) | 0.425 |
| period | Second period | 1,664 |  |  |
|  | First period | 2,137 | 1.043 (0.935-1.165) | 0.447 |
|  | Third period | 123 | 0.588 (0.407-0.874) | 0.006 |
| Interaction PERIOD and AGE | First period * 56-69 | - | 0.965 (0.845-1.1) | 0.591 |
|  | First period * 70-79 | - | 0.768 (0.657-0.897) | 0.001 |
|  | First period * >79 | - | 0.761 (0.508-1.136) | 0.182 |
|  | Third period * 56-69 | - | 1.401 (0.885-2.182) | 0.141 |
|  | Third period * 70-79 | - | 1.103 (0.682-1.76) | 0.683 |
|  | Third period * >79 | - | 1.438 (0.776-2.657) | 0.245 |
